# Social media use and internalising symptoms in clinical and community adolescent samples: a systematic review and meta-analysis

**DOI:** 10.1101/2023.09.12.23295355

**Authors:** Luisa Fassi, Kirsten Thomas, Douglas A Parry, Amelia Leyland-Craggs, Tamsin J Ford, Amy Orben

## Abstract

**Importance:** There are many concerns about the link between social media use and adolescent mental health. However, most research has studied adolescents from the general population, overlooking clinical groups. To address this gap, we synthesize, quantify and compare evidence on the relationship between social media use and internalising symptoms in adolescent clinical and community samples.

**Data Sources:** We searched five electronic databases for peer-reviewed publications (MEDLINE, Web of Science, PsychInfo, Scopus) and preprints (Europe PMC) published in English between 2007 and 2022.

**Study Selection:** We included cross-sectional and longitudinal studies quantifying the relationship between social media use and internalising symptoms, excluding experimental studies and randomised controlled trials. Two blinded reviewers initially identified 7389 studies.

**Data Extraction and Synthesis:** We adhered to the PRISMA and MOOSE guidelines for selection and reporting. The data was pooled using a random-effect model and robust variance estimation. Two reviewers independently assessed the quality of evidence with the Quality of Survey Studies in Psychology Checklist.

**Main Outcomes and Measures:** We preregistered our hypotheses and primary study outcomes on PROSPERO (CRD42022321473). Articles were included if they reported at least one quantitative measure of a) social media use: time spent, active vs. passive use, activity, content, user perception, and other; and b) internalising symptoms: anxiety, depression or both.

**Results:** We reviewed 127 studies including 1,061,293 adolescents and 775 effect sizes, of which only 8% examined clinical samples. In these samples, we found a positive, significant yet small meta-correlation between social media use and internalising symptoms, both for time spent (*N* = 2893, *r* = .08, *95% CI* = [.01, .15], *p* = .033, *I*^*2*^ = 57.83) and user engagement (N = 859; *r* = .12, *95% CI* = [.09, .15], *p* = .002, *I*^*2*^ = 82.67). These associations mirrored those in community samples.

**Conclusions and Relevance:** We highlight a lack of research on clinical populations, a critical gap considering that public concerns centre on the increase in youth mental health symptoms at clinical levels. This paucity of evidence not only restricts the generalizability of existing research but also hinders our ability to effectively evaluate and compare the link between social media use and youth mental health.

---

Mental health in adolescents (defined as those aged 10-24 years)^1^ has declined substantially in recent years. The proportion of UK adolescents with a probable mental health condition has increased from 10% in 2017 to 25% in 2022.^2,3^ Globally, it is now estimated that one in five children and adolescents are affected by mental health conditions, most commonly internalising disorders such as anxiety and depression.^4^ The impact of these conditions is wide-reaching and long-lasting, affecting school attendance, interpersonal relationships, employment prospects, physical health and risk of suicide, the second leading cause of death among 15-29-year-olds worldwide.^4^ Many have raised concerns that social media, which has become ubiquitous among young people (97% are daily users),^5^ might be a factor accelerating current mental health declines.^6,7^

Scientific investigations attempting to map how social media use relates to adolescent mental health have failed to provide much clarity. While converging evidence has established a small negative cross-sectional relationship between time spent on social media and well-being,^8–11^ studies employing longitudinal designs, measuring social media use beyond “time spent” or quantifying mental health beyond general well-being have shown diverging results. Consequently, there remains much conflicting evidence about the relationship between social media use and anxiety or depression.^12–14^

To find the source of this heterogeneity, researchers have studied a range of individual differences that could moderate the link between social media use and mental health.^15–18^ While many studies have examined factors such as age, gender, or ethnicity, little attention has been given to the types of samples recruited, especially regarding their mental health status. Studies investigating social media use and mental health routinely recruit adolescents from the general population through schools, universities, or as part of nationally representative samples.^12–14^ Despite these samples can include participants experiencing mental health symptoms at clinical levels, they often fail to draw a distinction between these participants and those experiencing symptoms at sub-clinical or non-clinical levels.

Individuals with mental health conditions face unique challenges, such as isolation, stigma and educational disruption.^19^ Thus, the failure to account for the nature and severity of mental health symptoms in the examined sample severely restricts our ability to draw accurate inferences about the relationship between social media use and mental health. For instance, adolescents with anxiety disorders face heightened sensitivity to social comparison and fear of negative evaluation,^20,21^ which could lead them to use or be impacted by social media differently compared to their peers.

We address the extent and impact of this oversight in three steps. First, we complete a pre-registered systematic review to quantify the proportion of studies that investigate the relationship between social media use and internalising symptoms in adolescent clinical samples compared to community or non-clinical samples. Second, we perform a meta-analysis to calculate the pooled relationship between social media use (differentiating between “time spent” as well as other measures of social media engagement such as activities or user perception) and internalising symptoms in clinical samples. Third, we compare the strength and direction of this relationship across clinical and community samples, testing whether sample type moderates the link between social media use and internalising symptoms.

This work allows us to gauge whether and how current research in this area of substantial scientific and public interest can be used to make clinically informative recommendations. It also complements pre-existing qualitative reviews of the clinical literature^22^ by providing a quantitative synthesis of effect sizes in clinical populations, and a direct comparison with those found in community samples. Overall, these findings have the potential to inform academics, by identifying knowledge gaps to direct future research; clinicians, by summarising research studying relevant populations; and policymakers, by guiding evidence-based decision-making.

## Methods

### Search strategy

The review and meta-analysis protocol was preregistered with the International Prospective Register of Systematic Reviews (Prospero; CRD42022321473)^23^ on 6^th^ April 2022, following the PRISMA 2020 checklist (see OSF).^24^ A health-science librarian developed the search strategy in collaboration with LF. We searched MEDLINE, Web of Science, PsychInfo, and Scopus databases in May 2022 using a combination of controlled vocabulary and keywords applicable to each individual database (Supplementary, Section 1). The search string included three clusters, with at least one matching term for each. The first cluster included terms relating to social media use (e.g., social media OR social networking sites), the second cluster related to the investigated population (e.g., adolescents OR youth), and the third cluster referred to mental health outcomes (e.g., internalising symptoms OR anxiety OR depression). Forwards and backward citation tracing was conducted via Google Scholar in January/February 2023 to identify studies missed by the electronic search strategy. Given the rapidly developing nature of this field, we also searched for preprints on Europe PMC at this time.^25^

### Selection criteria

Articles retrieved in the search were selected for inclusion based on 1) publication and design; 2) social media use measures; 3) mental health measures and 4) population. First, we selected peer-reviewed articles and preprints published in English on or after January 2007, the first year that Facebook became available to anyone over 13 years old. We included cross-sectional and longitudinal studies testing the relationship between social media use and internalising symptoms, regardless of the effect size reported. We excluded experimental studies and randomised controlled trials.

Second, we included articles capturing social media use using time-based (self-reported and logged) or engagement-based measures. These were pre-defined in our protocol and classified into five categories: active vs passive use, activity, content, user perception, and other (Supplementary, Section 2, Table 1S). Studies needed to report at least one quantitative social media use measurement (i.e., using an interval or continuous scale) either across multiple or single platforms (Supplementary, Section 2). Given our interest in studying regular social media use, we excluded articles sampling only problematic social media use, social media addiction or its use in a specific context (e.g., during driving).

**Table 1.** Moderation analyses. Results of moderation analyses for the meta-correlation between internalising symptoms and A) time spent on social media and B) engagement-based social media use measures.

| Moderator | Level | n | k | Estimate | SE | t-value | 95% CI | p-value |
| --- | --- | --- | --- | --- | --- | --- | --- | --- |
| **A. Time spent on social media and internalising symptoms** |  |  |  |  |  |  |  |  |
| *Sample type* | Clinical (ref) | 7 | 15 |  |  |  |  |  |
|  | Community | 44 | 92 | 0.05 | 0.03 | 1.6 | - 0.02, 0.12 | .153 |
| *COVID-19* | Before (ref) | 42 | 97 |  |  |  |  |  |
|  | During | 7 | 11 | 0.05 | 0.04 | 1.15 | - 0.05, 0.14 | .282 |
| *Mental health measure* | Internalising (ref) | 4 | 5 |  |  |  |  |  |
|  | Depression | 44 | 65 | - 0.07 | 0.08 | -0.82 | - 0.30, 0.17 | .460 |
|  | Anxiety | 16 | 37 | - 0.07 | 0.08 | -0.82 | - 0.27, 0.13 | .440 |
| **B. Social media engagement and internalising symptoms** |  |  |  |  |  |  |  |  |
| *Sample type* | Clinical (ref) | 4 | 19 |  |  |  |  |  |
|  | Community | 55 | 199 | 0.03 | 0.02 | 1.60 | - 0.03, 0.10 | .210 |
| *COVID-19* | Before (ref) | 49 | 188 |  |  |  |  |  |
|  | During | 6 | 22 | -0.01 | 0.06 | -0.2 | - 0.15, 0.13 | .838 |
| *Mental health measure* | Internalising (ref) | 2 | 7 |  |  |  |  |  |
|  | Depression | 50 | 157 | 0.01 | 0.02 | 0.67 | - 0.20, 0.23 | .616 |
|  | Anxiety | 21 | 54 | 0.04 | 0.04 | 1.20 | - 0.22, 0.31 | .402 |
| *Social media measure* | Other (ref) | 10 | 18 |  |  |  |  |  |
|  | Active vs passive | 10 | 31 | 0.02 | 0.05 | 0.33 | - 0.08, 0.11 | .744 |
|  | Activity | 19 | 65 | 0.09 | 0.06 | 1.60 | - 0.30, 0.21 | .132 |
|  | Content | 2 | 25 | - 0.07 | 0.04 | - 1.57 | - 0.35, 0.21 | .300 |
|  | Perception | 29 | 99 | 0.11 | 0.05 | 2.21 | 0.00, 0.22 | **.047\*** |

Note. *n* refers to the number of studies while *k* refers to the number of effect sizes for each level of the considered moderator.
\* $p < .05$

We included studies using quantitative clinical or symptom-based questionnaires to measure anxiety, depression, or both (Supplementary, Section 3, Table 2S). The latter, referred to as internalising symptoms, were included if they explicitly examined symptoms of both depressive and anxiety disorders in one outcome measure. We excluded articles measuring outcomes such as general health, psychopathology, well-being or life satisfaction.

Articles were included if they tested adolescents, defined as those between 10-24 years.^1^ If the age range was not provided, we required the mean age ± 1 SD to fall within the predetermined age range. Samples were categorised as clinical, community or non-clinical. We operationalised clinical samples as those including adolescents meeting any of the following criteria: 1) scoring above a clinical threshold on a symptom-based clinical questionnaire, 2) reporting an active clinical diagnosis, 3) accessing mental health services or being psychiatrically hospitalised. Community samples were defined as those including adolescents across the entire distribution of internalising symptoms without separation into clinical and non-clinical levels, while non-clinical samples were identified as those that specifically excluded adolescents in the clinical range.

We did not include studies that only tested clinical populations with symptomatology unrelated to internalising conditions (e.g., externalising symptoms or neurodevelopmental conditions). However, in line with transdiagnostic research that stresses the greater validity of accounting for comorbidities, we included studies of clinical samples with internalising conditions and comorbidity with other mental health conditions.^26^ If a study of a community sample reported two separate coefficients for adolescents scoring above and below clinical thresholds, we included clinical and non-clinical effect sizes separately. If studies reported including adolescents above clinical thresholds but failed to provide separate effect sizes, we requested these from the authors (Supplementary, Section 4).

### Study selection

We initially identified 12,984 records (7,770 articles and 5,214 preprints; Figure 1). All citations were saved on the Zotero reference management software and uploaded to Rayyan by two reviewers (LF and KT).^27^ Records were reduced to 7,389 (4,286 articles and 3,103 preprints) by removing duplicates. In a double-blinded process, LF and KT screened the title and abstract of the first 10% of articles (*N* = 428) applying the inclusion/exclusion criteria to decide whether to include, exclude or “maybe include”. Reviewers disagreed for <1% (5/428) of cases; these, as well as cases of “maybe include” (36/428), were discussed collaboratively. Afterwards, both reviewers screened the remaining 90% of articles independently, resolving disagreement (52/3852) and cases of “maybe include” (42/3852) through discussion.

**Figure 1.**
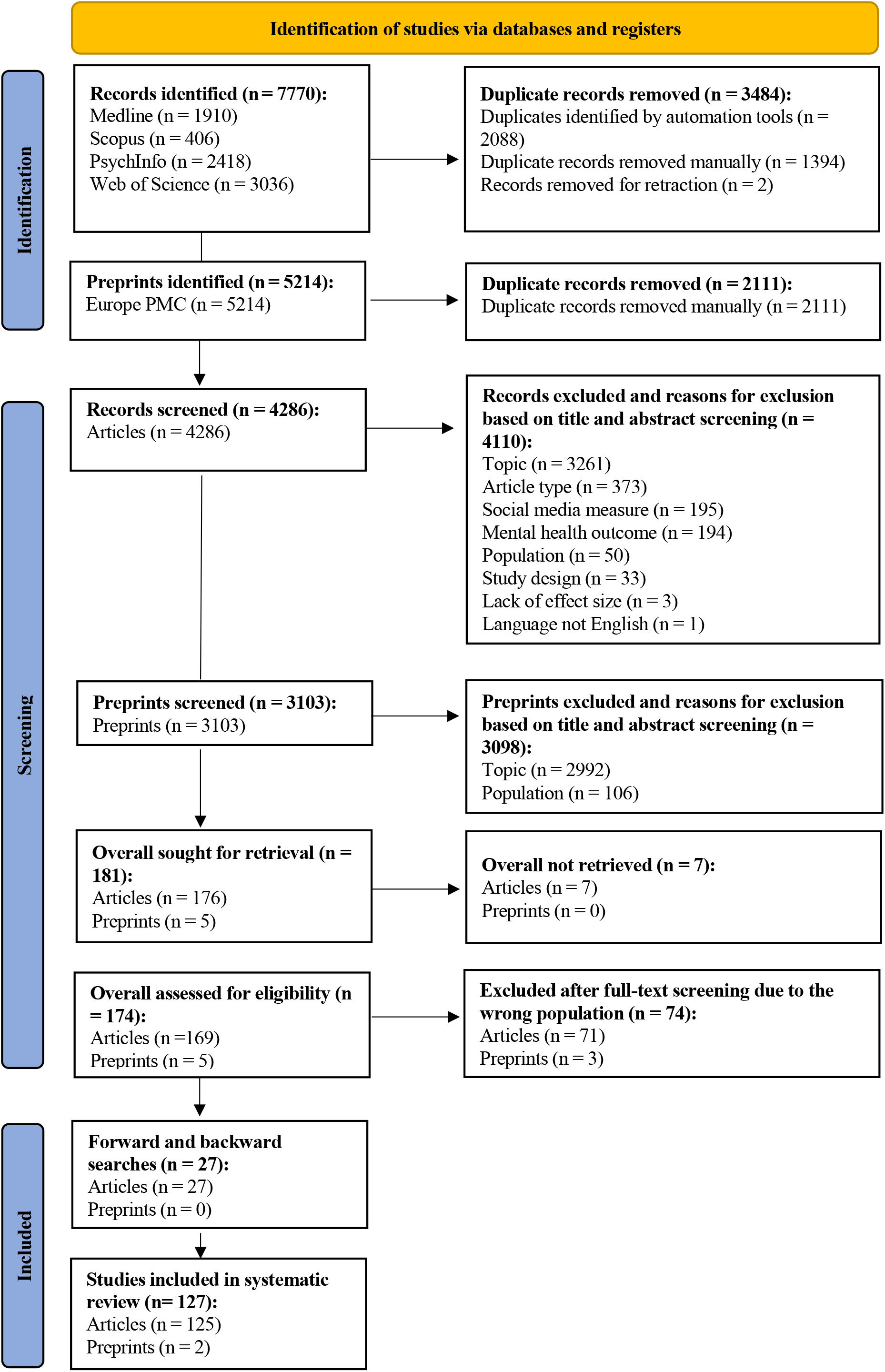
Flow diagram for study selection.

### Data extraction and quality assessment

Our standardised data extraction form included study information, participant characteristics, sample type (clinical, community, non-clinical), mental health measure (depression, anxiety, internalising), social media measure (time, active vs passive, activity, content, user perception, other), social media data collection methodology (self reported, logged), study design (cross-sectional, longitudinal), global population (global north, global south), and information to calculate pooled estimates of the relationship between social media use and mental health. Coders LF, ALC and KT independently coded the first 10% of articles and discussed any discrepancies to harmonise the coding strategy. Thereafter, coders KT and ALC coded 60% of the remaining papers each. The 20% of papers coded by both were used to calculate reliability (95%). All discrepancies were resolved through a discussion moderated by LF.

KT and ALC independently assessed the risk of bias and quality of included studies using the quality of survey studies in psychology (Q-SSP) checklist,^28^ resolving disagreements through discussion moderated by LF. This checklist was developed to evaluate the quality of cross-sectional and longitudinal psychological survey studies using 20 items. We adapted it to account for the type of evidence, such as secondary data, included in this study (Supplementary, Section 10).

### Data analysis

We completed all analyses in the R statistical programming language (Version 4.1.2); the full list of packages used can be found in the code shared on the OSF.^29^ We first conducted descriptive analyses of the studies included in the systematic review. Specifically, we calculated the number of studies and effect sizes (with associated percentages), split by sample type, mental health measure, social media measure, social media data collection methodology, study design and global population. Next, we conducted meta-analyses to test the pooled relationship between social media use and internalising symptoms for clinical samples, as well as community samples. The relationship was defined as positive when increased social media use was associated with increased internalising symptoms. We used an a priori statistical significance level of *α* = 0.05 and interpreted effect sizes in line with Cohen (1988; small effect: correlation coefficient of <0.10, medium effect: 0.30, large effect: >0.50). For studies reporting effect sizes other than a correlation coefficient, we performed transformations where possible (Supplementary, Section 5). We transformed all correlations from Fisher’s z back to Pearson’s r for reporting.

We used a random-effects model to calculate summary effect sizes due to the expected high level of heterogeneity. To account for variance inflation emerging from dependent observations for different measures collected from the same participants, we employed cluster-robust variance estimation (RVE) based on the sandwich method with adjusted estimators for small samples and correlated effects weighting scheme using the ‘robumeta’ package.^30–32^ We used a default value of *r* = .80 for the within-study effect size correlation. Sensitivity analyses showed that using different *r* values did not affect the inferences made about the overall effect size (see code shared on OSF).^29^ Given that longitudinal studies have multiple waves per participant, we included only the effect size from the first wave in the meta-analysis, to minimise variance inflation. Sensitivity analyses showed no differences in the strength and direction of the meta-analytic effect sizes if we included all waves (Supplementary, Section 9).

Our confirmatory meta-analyses examining the relationship between social media use and internalising symptoms were restricted to studies measuring time spent on social media, to allow meaningful pooling of effect sizes across studies due to measurement similarity. If a study measured social media use as time spent and reported effect sizes for overall time spent as well as specific time spent on individual applications (e.g., Instagram and Twitter), we excluded the latter to avoid nested correlations. However, if no measure of overall time spent was available, we included separate effect sizes for time spent on individual applications. We also conducted exploratory meta-analyses focusing on engagement-based social media use measures (Supplementary, Section 2).

### Risk of bias assessment and moderation

To assess potential bias due to small-study effects, including publication bias, we visually inspected funnel plot symmetry and performed Egger’s regression test.^33,34^ Further, we used a contour-enhanced funnel plot with superimposed areas of statistical significance (corresponding to *p* = .10, .05 and .01), interpreting an over-representation of effect sizes in the highlighted areas as indicative of potential publication bias.^34^ We conducted influence diagnostics (Cook’s distance, covariance ratios and diagonal elements of the hat matrix) using the ‘metafor’ package^35^ to determine whether analyses were impacted by outliers and performed leave-one-out sensitivity analyses with such outliers removed.

To examine heterogeneity in effect sizes, we computed *I*^*2*^, interpreting values of approximately 25%, 50%, and 75% to indicate low, moderate, and high heterogeneity, respectively. We conducted three pre-registered moderator analyses to investigate factors contributing to heterogeneity in the pooled correlation between social media use and internalising symptoms. This included sample type (comparing clinical and community samples; non-clinical samples were excluded due to a lack of power), mental health measure (anxiety, depresssion or internalising symptoms) and COVID-19 (before, during), given its impact on adolescents’ lifestyle,^36^ mental health^37^ and technology use.^38^ We classified studies as happening during the pandemic if any data collection was performed after January 2020.^4^

## Results

### Systematic review: Quantifying the proportion of clinical samples

After duplicate removal, we screened 7,389 manuscripts (4,286 articles and 3,103 preprints), including 127 studies in the systematic review (125 articles and 2 preprints, Figure 1). Included studies had a combined sample size of *N* = 1,061,293 adolescents (*m* = 8,356; *sd* = 42,424; *mdn* = 680, *min* = 41, *max* = 388,275) and reported 775 effect sizes for the relationship between social media use and internalising symptoms.

Studies investigating adolescent clinical samples were rare: 8% of effect sizes, corresponding to 59 effect sizes from 11 studies (Figure 2A and Supplementary Section 11, Table 3S for a qualitative summary of these studies).

**Figure 2.**
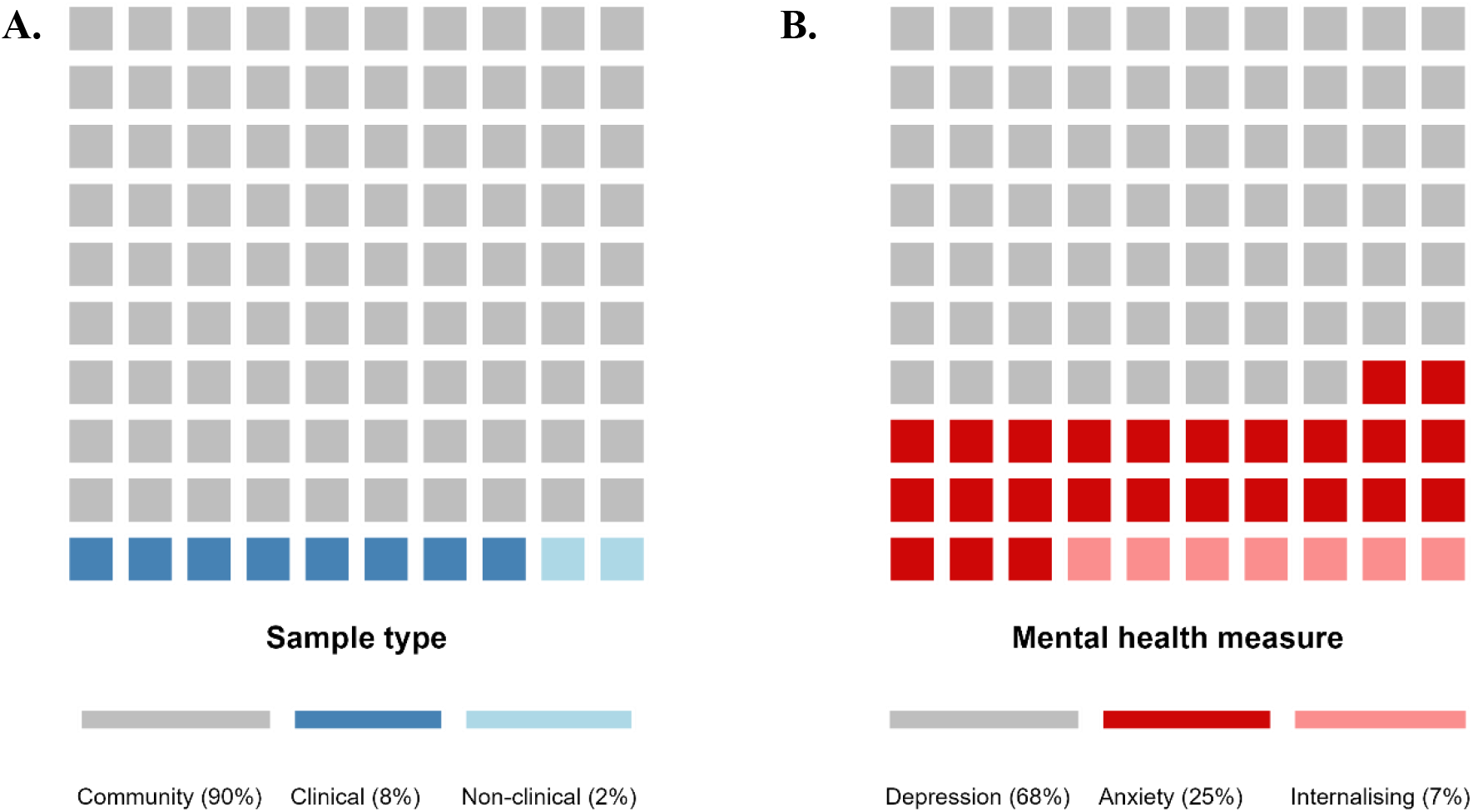
Proportion of included effect sizes by sample type and mental health measure. Grid of 10x10 (100%) squares representing the percentage of literature in the systematic review by A) sample type and B) mental health measure. The presented proportion is calculated based on the total number of effect sizes (N = 775). *Note*. We used the total number of effect sizes instead of the total number of studies to calculate proportions given that an individual study could include more than one sample type or mental health measure.

The vast majority of studies examined community samples (90% of effect sizes; 703 effect sizes from 117 studies), with very few studying non-clinical samples (2% of effect sizes, 13 effect sizes from 4 studies). The most common mental health outcome studied was depression (68% of effect sizes; 523 effect sizes from 105 studies); while anxiety (25% of effect sizes; 194 effect sizes from 45 studies) and internalising symptoms (7% of effect sizes; 58 effect sizes from 14 studies) were less frequently assessed (Figure 2B).

Further, 92% of effect sizes were derived from studies using self-reported social media use measures (711 effect sizes from 122 studies), as only 8% used objective logged measures (64 effect sizes from 6 studies). Nearly half of effect sizes were extracted from studies measuring time spent on social media (47%, 366 effect sizes from 81 studies). Less common engagement-based measures of social media use included user perception (20%, 157 effect sizes from 34 studies), activity (16%, 125 effect sizes from 27 studies), active versus passive use (6%, 42 effect sizes from 10 studies), content (4%, 29 effect sizes from 4 studies), and other metrics (7%, 56 effect sizes from 18 studies). Most studies (65%, 82 studies) were cross-sectional, while 35% (45 studies) were longitudinal. In line with previous work,^16^ the most commonly studied populations were from the Global North (83%; 106 studies), compared to the Global South (17%; 21 studies).

Overall, approximately half of the included studies were of acceptable quality (67/127 studies, 52.76%, compared to the remaining 47.24% that were classified as being of questionable quality; Supplementary, Section 10).

### Meta-analysis: quantifying links in clinical samples

#### Social media time spent

Seven studies of clinical populations (15 effect sizes) used measures of time spent on social media. The total sample size was *n* = 2,893 (*m =* 413; *sd* = 585; *mdn* = 224; *min* = 49; *max* = 1,722). In our confirmatory meta-analysis, we found a positive, significant, yet small meta-correlation between time spent on social media and internalising symptoms (Figure 3, *r* = .08, 95% *CI* = [.01, .15], *p* = .033), with moderate heterogeneity (*I*^*2*^ = 57.83). After removing one outlier (*ID* = P155E19), the meta-correlation coefficient increased slightly (*r* = .10, 95% *CI* = [.02, .18], *p* = .023), while heterogeneity decreased (*I*^*2*^ = 26.56). Further, Egger’s regression test showed no evidence of small study bias (*β* = -2.19, *se* = 0.46, *p* = .984; also confirmed by visual inspection of the funnel plot; Supplementary, Section 7, Figure S2C-D).

**Figure 3.**
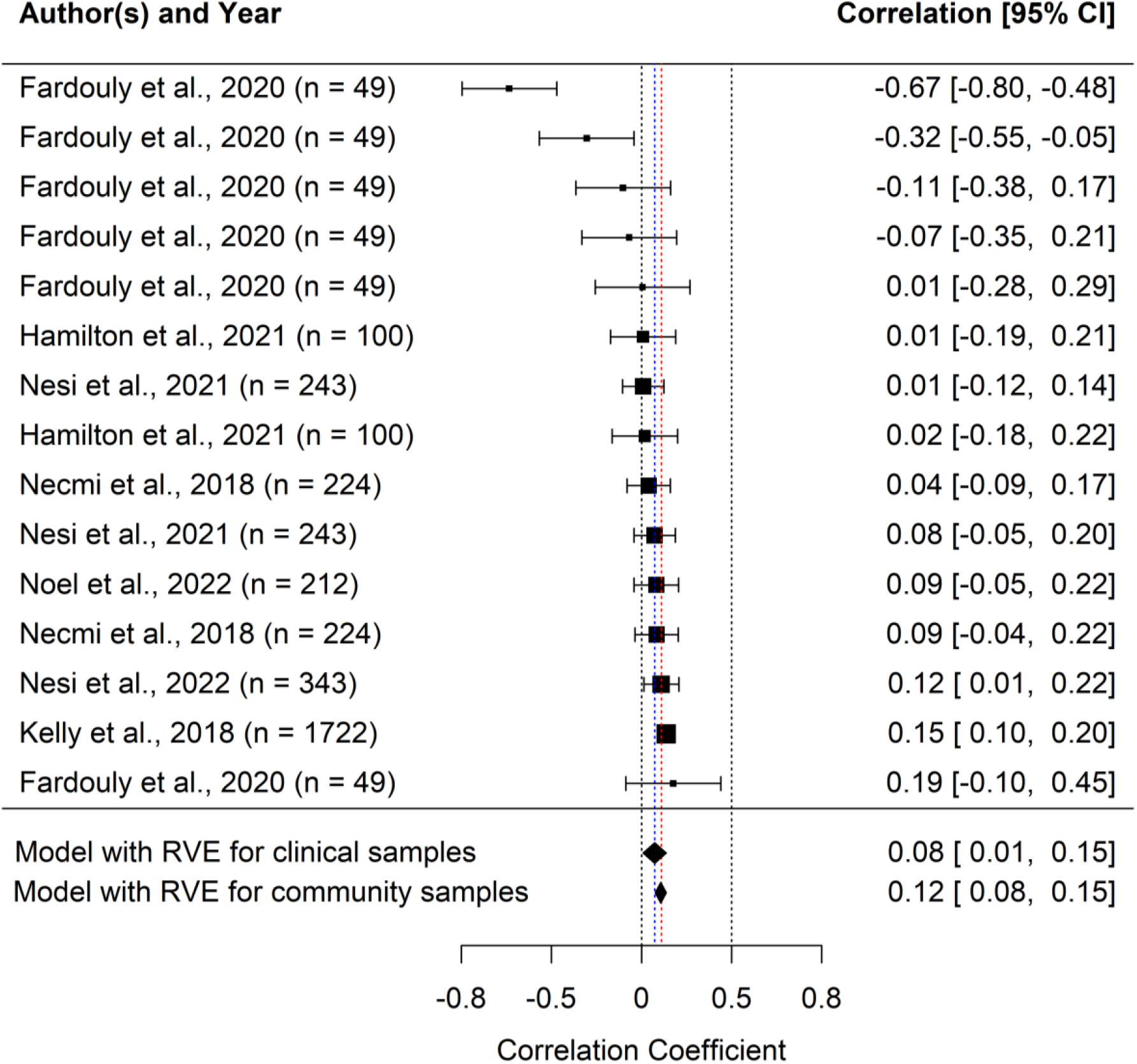
Forest plot for time spent on social media in clinical groups. Forest plot for the individual and pooled effect sizes representing the relationship between time spent on social media and internalising symptoms. Effect sizes for clinical samples are shown both individually (i.e., separate rows with author and year) and as a pooled estimate (model with RVE), while the effect size for community samples is only presented as a pooled estimate at the bottom. *Note*. Individual Pearson’s r coefficients are depicted as filled squares with the size indicating the relative weight, based on sample size, of each effect size estimate for clinical studies in the meta-analysis. Increased time spent on social media is associated with decreased internalising symptoms (to the left of zero) or increased internalising symptoms (to the right of zero). The first filled black diamond and the dashed blue line represent the overall summary effect size across all clinical studies (*r* = .08, *95% CI* = [.01, .15], *p* < .001), calculated using robust variance estimation to account for dependencies between effect sizes coming from the same study. The second filled-back diamond and the dashed red line represent the overall summary effect size across all studies run on community samples (*r* = .12, *95% CI* = [.08, .15], *p* < .001). The error bars and diamond width represent the 95% CIs for the effect sizes. The dashed reference line at *r* = 0 represents the point of reference for no correlation, while the dashed reference line at the intercept for *r* = 0.5 represents the point from which the magnitude of the correlation would be sufficient to conclude that there is a moderate relationship between time spent on social media and internalising symptoms.

#### Social media engagement

The need to move beyond “time spent” measures of social media use has been widely acknowledged, as these measures are simplistic and fail to distinguish between types of activities or content that can differentially impact mental health.^39,40^ Researchers have therefore advocated for using engagement-based measures of social media use, which we examined in an exploratory meta-analysis. Four studies of clinical populations (19 effect sizes) used engagement-based measures of social media use, specifically social media activities (10 effect sizes) and user perception (9 effect sizes), with a total sample size of *n* = 859 (*m =* 215; *sd* = 122; *mdn* = 233; *min* = 49; *max* = 343). We found a positive and significant, yet small meta-correlation between these social media measures and internalising symptoms (Figure 4, *r* = .12, 95% *CI* = [.09, .15], *p* = .002), with high heterogeneity (*I*^*2*^ = 82.67). No outliers were identified. Further, Egger’s regression test showed no evidence of small study bias (*β* = -0.55, *se* = 0.15, *p* = .928; also confirmed by visual inspection of funnel plot; Supplementary 7, Figure S4C).

**Figure 4.**
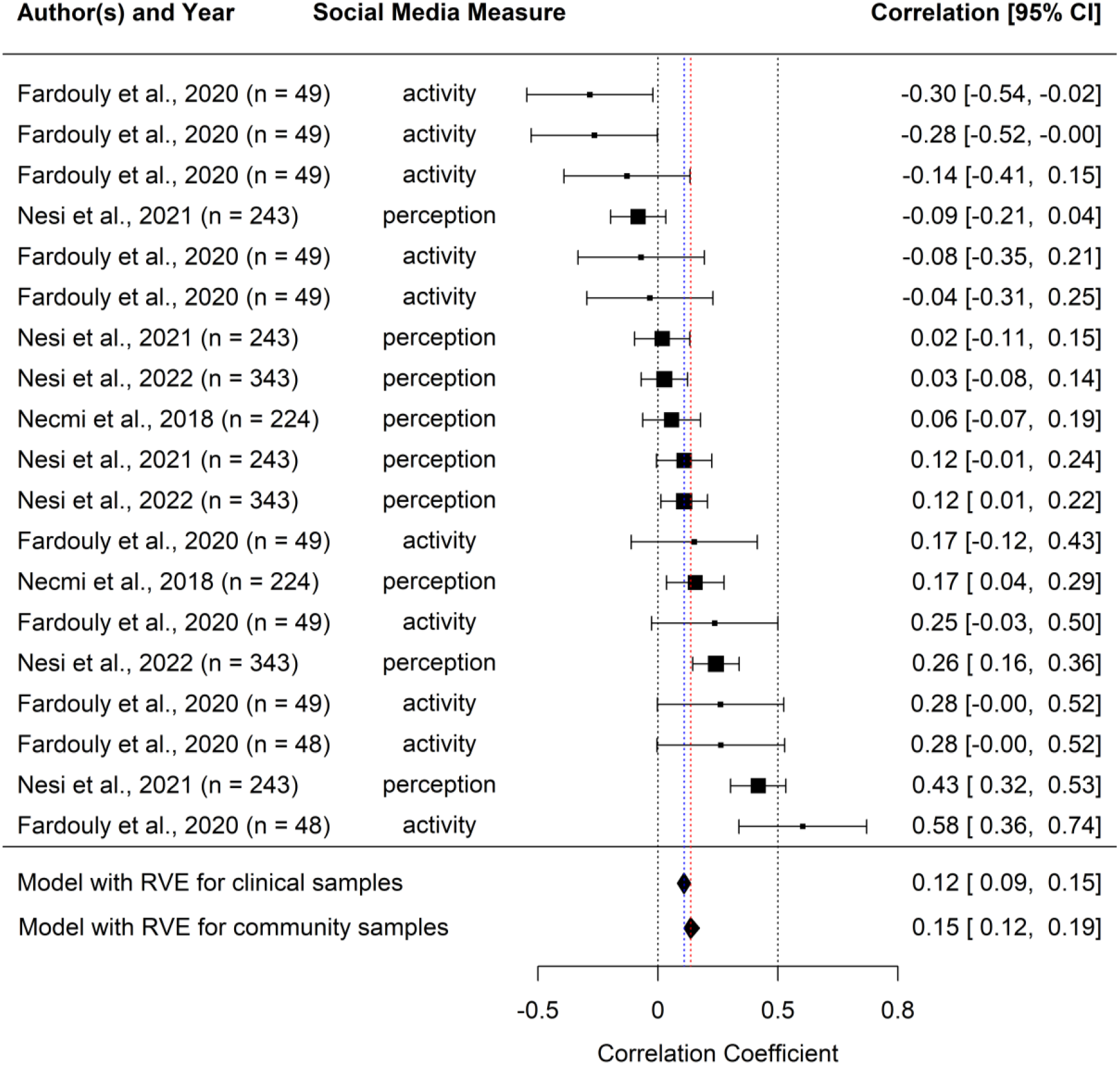
Forest plot for social media engagement in clinical groups. Forest plot for the individual and pooled effect sizes representing the relationship between social media engagement and internalising symptoms. Effect sizes for clinical samples are shown both individually (i.e., separate rows with author and year) and as a pooled estimate (model with RVE), while the effect size for community samples is only presented as a pooled estimate at the bottom. *Note*. Individual Pearson’s r coefficients are depicted as filled squares with the size indicating the relative weight, based on sample size, of each effect size estimate for clinical studies in the meta-analysis. Increased social media engagement is associated with decreased internalising symptoms (to the left of zero) or increased internalising symptoms (to the right of zero). The first filled black diamond and the dashed blue line represent the overall summary effect size across all clinical studies (*r* = .11, *95% CI* = [.06, .16], *p* < .001), calculated using robust variance estimation to account for dependencies between effect sizes coming from the same study. The error bars and diamond width represent the 95% CIs. The second filled back diamond and the dashed red line represent the overall summary effect size in community samples (*r* = .15, *95% CI* = [.12, .19], *p* < .001). The dashed black reference line at *r* = 0 represents the point of reference for no correlation, while the dashed reference line at the intercept for *r* = 0.5 represents the point from which the magnitude of the association would be sufficient to conclude that there is a moderate relationship.

### Meta-analysis: comparing links between clinical and community samples

#### Social media time spent

We also ran a meta-analysis of the 44 studies (and 92 effect sizes) testing community samples that were identified by our systematic review (*n* = 469,230; *m =* 10,664; *sd* = 58,540; *mdn* = 460; *min* = 41; *max* = 388,275). We found a positive and significant, yet small meta-correlation between time spent on social media and internalising symptoms (*r* = 0.12, 95% *CI* = [0.08, 0.15], *p* < .001; see Figure 3). This is similar to the meta-correlation found in clinical samples (*r* = 0.08, 95% *CI* = [0.01, 0.15], *p* = .033, *I*^*2*^ = 57.83) but shows higher levels of heterogeneity: *I*^*2*^ = 98.40 (Supplementary, Section 8, Figure S3 for the distribution of effect sizes).

To further test whether sample type (clinical vs community) influences the link between time spent on social media and internalising symptoms, we ran a combined meta-analysis (51 studies with 110 effect sizes; *n* = 472,288; *m =* 9,260; *sd* = 54,404; *mdn* = 407; *min* = 41; *max* = 388,275) and tested sample type as a moderator. We found an overall positive yet small meta-correlation between social media use and internalising symptoms across all sample types (*r* = 0.11, *95% CI* = [.08, .14], *p* < .001). This finding was robust after one outlier was removed (*ID* = P155E19, *r* = 0.11, *95% CI* = [.09, .14], *p* < .001). Heterogeneity was high in both cases (*I*^*2*^ = 98.14) and there was no evidence of small study bias (RVE for the Egger–sandwich test: *β* = - 0.93, *se* = 0.51, *p* = 0.956; also confirmed by visual inspection of funnel plot; Supplementary, Section 7, Figure S2A-B).

After excluding non-clinical samples due to a lack of power (3 effect sizes from 3 studies), we tested sample type as a moderator, with two levels: clinical vs community sample. We found non-significant results (Table 1A, *β* = 0.05, *se* = 0.03, *t* = 1.6, *95% CI* = [-0.02, 0.12], *p* = .153), with high heterogeneity (*I*^*2*^ = 98.22). Results did not change after removing one outlier (*n* = P155E19; *β* = 0.04, *se* = 0.03, *t* = 1.52, 95% *CI* = [-0.02, 0.10], *p* = .172).

Sample type is therefore not a key factor explaining differences in the relationship between time spent on social media and adolescents’ internalising symptoms.

We also tested whether mental health measures (three levels: anxiety, depression, internalising) and COVID-19 (two levels: before vs. during) were moderators.

Yet, neither mental health measure (depression vs internalising, *β* = - 0.07, *se* = 0.08, *t* = - 0.82, *95% CI* = [-0.30, 0.17], *p* = .460; anxiety vs internalising, *β* = - 0.07, *se* = 0.08, *t* = - 0.82, *95% CI* = [-0.27, 0.13], *p* = .440) nor COVID-19 (*β* = 0.05, *se* = 0.04, *t* = 1.15, *95% CI* = [-0.05, 0.14], *p* = .282) explained heterogeneity in the meta-correlation between time spent on social media and internalising symptoms (Table 1A).

#### Social media engagement

We repeated the same analyses for studies measuring social media engagement. As with the meta-correlation of studies using clinical samples (*r* = .12, 95% *CI* = [.09, .15], *p* = .002, *I*^*2*^ = 82.67), we found a small, positive, and significant meta-correlation between social media engagement and internalising symptoms (*r* = .15, 95% *CI* = [.12, .19], *p* < .001; see Figure 4) in studies run on community samples (199 effect sizes from 55 studies, *n* = 61,294; *m =* 1,114; *sd* = 1,690; *mdn* = 551, *min* = 41, *max* = 10,563). There were high levels of heterogeneity (*I*^*2*^ = 94.91; Supplementary, Section 7 and 8, Figure S3B for the distribution of effect sizes) and results remained robust after removing one outlier (*ID* = P97E6, *r* = .15, 95% *CI* = [.12, .19], *p* < .001, *I*^*2*^ = 94.83).

We included 58 studies with 218 effect sizes in our combined meta-analysis. As before, non-clinical samples were excluded due to lack of power. Across all sample types *(n =* 64,302; *m =* 1,108; *sd* = 1,683; *mdn* = 551; *min* = 41; *max* = 10,563), there was a positive yet small meta-correlation between social media engagement and internalising symptoms (*r* = 0.15, *95% CI* = [0.12, 0.19], *p* < .001), with high heterogeneity (*I*^*2*^= 94.66). Results remained robust after removing one outlier (*ID* = P97E6, *r* = 0.15, *95% CI* = [0.12, 0.18], *p* < .001, *I*^*2*^= 94.58). There was no evidence of small study bias (*β* = 0.69, *se* = 0.66, *p* = 0.156 also confirmed by visual inspection of the funnel plot, Supplementary, Section 7, Figure S4A-B).

Sample type (clinical vs community) was not a significant moderator of the overall relationship between social media engagement and internalising symptoms (Table 1B, *β* = 0.03, *SE* = 0.02, *t* = 1.6, 95% *CI* = [-0.03, 0.10], *p* = .210), and heterogeneity remained high (*I*^*2*^ = 94.74). Our additional moderation analyses, summarised in Table 1B, showed that neither mental health measure (anxiety vs internalising, *β* = 0.04, *se* = 0.04, *t* = 1.20, *95% CI* = [-0.22, 0.31], *p* = .402.; depression vs internalising, *β* = 0.01, *se* = 0.02, t = 0.67, *95% CI* = [-0.20, 0.23], *p* = .616) nor COVID-19 (*β* = -0.01, *se* = 0.06, *t* = -0.21, *95% CI* = [-0.15, 0.13], *p* = .838) explained heterogeneity in the meta-correlation between social media engagement and internalising symptoms. There were also no moderator effects for different types of social media measures, except for user perception (*β* = 0.11, *se* = 0.05, *t* = 2.21, *95% CI* = [0.00, 0.22], *p* = .047).

## Discussion

This systematic review and meta-analysis synthesised data from 15 years of research examining the relationship between social media use and internalising symptoms in over one million adolescents. We found that only 8% of studies examined clinical populations, while 90% recruited adolescents from the general population. There was a small, positive, and significant meta-correlation between social media use and internalising mental health in clinical samples, regardless of whether time- or engagement-based social media metrics were studied. Notably, these meta-correlations did not substantially differ from those found in community samples. Indeed, sample type (clinical vs community) was not a significant moderator of the meta-correlation between social media use and adolescents’ internalising mental health.

There is therefore a critical lack of research on social media use and mental health in adolescent clinical populations. As these groups differ from the general population, this severely limits our capacity to draw accurate inferences about the relationship between social media use and mental health. For instance, adolescents experiencing anxiety and depressive disorders face social withdrawal, low self-esteem, increased susceptibility to peer influence, heightened sensitivity to feedback and excessive rumination,^41^ all of which may alter their interaction with social media, and its ultimate impact on their mental health.^42–44^

In contrast to the common assumption that clinical populations might show a stronger relationship between social media use and mental health declines than community samples,^45^ our findings indicate no substantial differences. This could be because the mental health status of the examined sample is not a key factor explaining group-level differences. For example, adolescents might adjust their social media use based on their mental health needs, leading to comparable usage patterns and correlations. However, clinical populations could also be experiencing less variability in mental health symptomatology (e.g., ceiling effects), lessening the observable correlations between mental health symptoms and social media use. Further, common methodological limitations, such as inaccurate self-report measures of social media use,^46^ might decrease our ability to locate differences even if they exist. While our work represents the most comprehensive synthesis of the available evidence in clinical samples to date, the lack of research on these populations additionally constrains our ability to draw accurate comparisons.

We also underscore some additional limitations. Studies included in our meta-analysis captured the relationship between social media use and internalising mental health using correlations. Hence, no causal inferences can be drawn about whether increased social media use leads to internalising symptoms or vice versa. In addition, we focused on internalising mental health only, and inferences cannot be generalised to other clinical groups (e.g., externalising conditions). We also categorised social media engagement using five pre-defined categories, which are not exhaustive and could mask important differences in social media engagement. For example, the impact of social media content will depend on its nature, which could be positive, negative or neutral. Finally, our meta-analyses demonstrated the moderate to high levels of heterogeneity common to this research area.^13^ While this variation could potentially be explained by individual differences such as demographics, this could not be tested due to a lack of statistical power.

Many worry about social media’s role in increased clinical-level mental health symptoms among adolescents. However, current research falls short of adequately targeting the specific populations required to draw accurate inferences about this matter. Despite our initial findings of a similar association across clinical and community samples, there is still a real risk that we are incorrectly generalising results from the general population to young people with mental health conditions. The potential impact of this extends far beyond research to clinical practice and policymaking. In a world increasingly saturated by digital technology, we cannot afford to design prevention programs, interventions, patient consultations, and regulations without knowing that they work for everyone, especially those most vulnerable.

## Data Availability

All data produced are available online at the Open Science Framework.

https://osf.io/94bpj/?view_only=6b1458debe3b4432be9679c65403a6f1

https://osf.io/gr3xh/?view_only=7b7434c46ad44d6fbbeafcd1481f2e93

## Preregistration and protocol

This study was preregistered on PROSPERO (CRD42022321473) and OSF (https://osf.io/fcvwy/?view_only=192a5df5aa2a4104b414c3d284e80906)

## Data and code availability

The data (https://osf.io/94bpj/?view_only=6b1458debe3b4432be9679c65403a6f1) and code (https://osf.io/gr3xh/?view_only=7b7434c46ad44d6fbbeafcd1481f2e93) are available on the OSF.

## Supplementary material

The supplementary material for this article can be found on OSF (https://osf.io/uwhrc/?view_only=c8508b4f8e8346fba9bf99c7bc44b346).

## Authors’ contributions

We present author contributions according to the CRediT (Contributor Roles Taxonomy).

L.F: Conceptualisation, methodology, formal analysis, writing - original draft, writing - review & editing.

K.T: Conceptualisation, methodology, writing - review & editing.

D.P: Conceptualisation, methodology, formal analysis, writing - review & editing, supervision.

A.L: Methodology, writing - review & editing.

T.F: Conceptualisation, writing - review & editing, supervision.

A.O: Conceptualisation, methodology, writing - review & editing, supervision.

