## Supplementary Materials for "Social media use and internalising symptoms in clinical and community adolescent samples: a systematic review and meta-analysis"

### Supplementary Information

#### Section 1: Search strings

We used the following search strings for the different databases: MEDLINE, PsychInfo, Web of Science, Scopus and Europe PMC.

*MEDLINE:*

1. (social-media or online-communit* or social-app* or social-networking-app* or social-networking-site or social-networking-sites or social adj3 (media or network* or application* or app* or network* or platform*) or online adj2 (communicat* or messag* or platform*) or Facebook or YouTube or WhatsApp or Messenger or Snapchat or WeChat or Instagram or QQ or Tumblr or Qzone or Tik-Tok or Sina-Weibo or Twitter or Reddit or Baidu or LinkedIn).ti,ab. or Social Media/ or Social Networking/
2. (adolescen* or teen* or youth* or ((young and (people or person* or adult*)) or juvenile or "high school student*" or "secondary school student*" or undergraduate or postgraduate)).ti,ab. or Adolescent Psychiatry/ or Adolescent Behavior/ or Adolescent, Hospitalized/ or Adolescent/ or Adolescent Health/ or Psychology, Adolescent/ or Adolescent Development/
3. ((mental or emotional or psychosocial) adj2 problem* or mental health adj2 (problem* or disorder* or risk* or poor) or (behaviour* or behavior*) adj2 (problem* or disorder* or risk*) or depress* or anxiety or "major depressive disorder" or stress* or distress* or "emotional health" or psychopathology or internali* adj2 (problem* or condition*)).ti,ab. or Depression/ or Anxiety/ or anxiety disorders/ or agoraphobia/ or anxiety, separation/ or obsessive-compulsive disorder/ or panic disorder/ or phobic disorders/ or "bipolar and related disorders"/ or mood disorders/ or depressive disorder/ or depressive disorder, major/ or depressive disorder, treatment-resistant/ or dysthymic disorder/ or premenstrual dysphoric disorder/ or seasonal affective disorder/
4. 1 AND 2 AND 3

Filters: Additional limits - Date: From January 2007; Language: English

*PsychInfo:*

1. TI (social-media or online-communit* or social-app* or social-networking-app* or social-networking-site or social-networking-sites or social n3 (media or network* or application* or app* or network* or platform*) or online n2 (communicat* or messag* or platform*) or Facebook or YouTube or WhatsApp or Messenger or Snapchat or WeChat or Instagram or QQ or Tumblr or Qzone or Tik-Tok or Sina-Weibo or Twitter or Reddit or Baidu or LinkedIn) or DE social media
2. AB((social-media or online-communit* or social-app* or social-networking-app* or social-networking-site or social-networking-sites or social n3 (media or network* or application* or app* or network* or platform*) or online n2 (communicat* or messag* or platform*) or Facebook or YouTube or WhatsApp or Messenger or Snapchat or WeChat or Instagram or QQ or Tumblr or Qzone or Tik-Tok or Sina-Weibo or Twitter or Reddit or Baidu or LinkedIn)
3. TI(adolescen* or teen* or youth* or ((young and (people or person* or adult*)) or juvenile or "high school student*" or "secondary school student*" or undergraduate or postgraduate))

4. AB(adolescen* or teen* or youth* or ((young and (people or person* or adult*)) or juvenile or "high school student*" or "secondary school student*" or undergraduate or postgraduate))
5. TI/AB((mental or emotional or psychosocial) n2 problem* or mental health n2 (problem* or disorder* or risk* or poor) or (behaviour* or behavior*) n2 (problem* or disorder* or risk*) or depress* or anxiety or "major depressive disorder" or stress* or distress* or "emotional health" or psychopathology or internali* n2 (problem* or condition*)) or DE anxiety or DE depression or DE mood disorders
6. AB((mental or emotional or psychosocial) n2 problem* or mental health n2 (problem* or disorder* or risk* or poor) or (behaviour* or behavior*) n2 (problem* or disorder* or risk*) or depress* or anxiety or "major depressive disorder" or stress* or distress* or "emotional health" or psychopathology or internali* n2 (problem* or condition*))
7. (1 or 2) AND (3 or 4) AND (5 or 6)

Filters: Additional limits - Date: From January 2007; Language: English

*Web of Science:*

1. (TI=(social-media or online-communit* or social-app* or social-networking-app* or social-networking-site or social-networking-sites or social near/3 (media or network* or application* or app* or network* or platform*) or online near/2 (communicat* or messag* or platform*) or Facebook or YouTube or WhatsApp or Messenger or Snapchat or WeChat or Instagram or QQ or Tumblr or Qzone or Tik-Tok or Sina-Weibo or Twitter or Reddit or Baidu or LinkedIn)
2. (AB=(social-media or online-communit* or social-app* or social-networking-app* or social-networking-site or social-networking-sites or social near/3 (media or network* or application* or app* or network* or platform*) or online near/2 (communicat* or messag* or platform*) or Facebook or YouTube or WhatsApp or Messenger or Snapchat or WeChat or Instagram or QQ or Tumblr or Qzone or Tik-Tok or Sina-Weibo or Twitter or Reddit or Baidu or LinkedIn)
3. (AB=(adolescen* or teen* or youth* or ((young and (people or person* or adult*)) or juvenile or "high school student*" or "secondary school student*" or undergraduate or postgraduate))
4. (TI=(adolescen* or teen* or youth* or ((young and (people or person* or adult*)) or juvenile or "high school student*" or "secondary school student*" or undergraduate or postgraduate))
5. (AB/TI=((mental or emotional or psychosocial) near/2 problem* or mental health near/2 (problem* or disorder* or risk* or poor) or (behaviour* or behavior*) near/2 (problem* or disorder* or risk*) or depress* or anxiety or "major depressive disorder" or stress* or distress* or "emotional health" or psychopathology or internali* near/2 (problem* or condition*))
6. (TI=((mental or emotional or psychosocial) near/2 problem* or mental health near/2 (problem* or disorder* or risk* or poor) or (behaviour* or behavior*) near/2 (problem* or disorder* or risk*) or depress* or anxiety or "major depressive disorder" or stress* or distress* or "emotional health" or psychopathology or internali* near/2 (problem* or condition*))
7. (1 or 2) AND (3 or 4) AND (5 or 6)

Filters: Additional limits - Date: From January 2007; Language: English

*Scopus:*

TITLE-ABS-KEY((social-media or online-communit* or social-app* or social-networking-app* or social-networking-site or social-networking-sites or social n3 (media or network* or application* or app* or network* or platform*) or online n2 (communicat* or messag* or platform*) or Facebook or YouTube or WhatsApp or Messenger or Snapchat or WeChat or Instagram or QQ or Tumblr or Qzone or Tik-Tok or Sina-Weibo or Twitter or Reddit or Baidu or LinkedIn) AND ((adolescen* or teen* or youth* or ((young and (people or person* or adult*)) or juvenile or "high school student*" or "secondary school student*" or undergraduate or postgraduate) AND (mental or emotional or psychosocial) n2 problem* or mental health n2 (problem* or disorder* or risk* or poor) or (behaviour* or behavior*) n2 (problem* or disorder* or risk*) or depress* or anxiety or "major depressive disorder" or stress* or distress* or "emotional health" or psychopathology or internali* n2 (problem* or condition)) AND ( LIMIT-TO ( LANGUAGE,"English" ) ) AND PUBYEAR AFT 2006

*Europe PMC:*

(social-media or online-communication or social-app or social-networking-app or social-networking-site or social-networking-sites or Facebook or YouTube or WhatsApp or Messenger or Snapchat or WeChat or Instagram or QQ or Tumblr or Qzone or Tik-Tok or Sina-Weibo or Twitter or Reddit or Baidu or LinkedIn) AND (adolescent or teen or youth or young people or young person or young adult or juvenile or "high school student" or "secondary school student" or undergraduate or postgraduate) AND ("mental problem" or "emotional problem" or "psychosocial problem" or "mental health problem" or disorder or "behavioural problem" or "behavioural disorder" or depression or anxiety or "major depressive disorder" or stress or distress or "emotional health" or psychopathology or internalizing or "internalizing problems")

Filters: Additional limits - Date: From January 2007; Language: English

**Section 2: Inclusion criteria for social media use**

For the inclusion criteria regarding social media use, we selected articles that measured overall social media use or use of any of the following platforms: Facebook, YouTube, WhatsApp, Messenger, WeChat, Instagram, QQ, Tumblr, Qzone, Tik-Tok, Sina Weibo, Twitter, Reddit, Baidu Tieba, LinkedIn, Viber, Snapchat, Pinterest, Line or Telegram. We categorised types of social media engagement based on five categories in our preregistered protocol. These categories were defined based on prior research that highlights the importance of distinguishing between various forms of social media engagement, beyond simply the time spent on a platform/social media in general.[1–3]

**Table 1S.**

Inclusion criteria and categorisation for social media measures.

| Category of social media measure | Definition and examples of measures | Use in systematic review/ meta-analysis |
|---|---|---|
| Time spent and frequency | This category includes:<br>1) measures of the amount of time spent on social media over a specified time period (e.g., self-reported or logged hours per day);<br>2) number/count of general social media usage behaviours over a specific time period (e.g., self-reported or logged number of log-ins or usage sessions per day).<br><br>This category does not include the amount of time/count of social media behaviours that are associated with a specific activity (e.g., posting *N* pictures) or content (e.g., looking at *N* posts with food content). These would be categorised as *activity* or *content* measures, respectively. | Systematic review and confirmatory meta-analysis |
| Activity | Time/frequency of a specific activity on social media (e.g., number of pictures posted, amount of time spent messaging). | Systematic review and exploratory meta-analysis |
| Content | Time/frequency of exposure on social media to specific content (e.g., amount of minutes in a day spent looking at food content, appearance-related content, inspirational content, self-harm content and other types of content). | Systematic review and exploratory meta-analysis |
| User perceptions | Quantification of users' motivations and attitudes about using social media, and reflections about their social media use (e.g., the Social Media Use Integration scale). | Systematic review and exploratory meta-analysis |
| Active vs passive use | Time/frequency of active or passive social media use (e.g., the Passive and Active Facebook Use Measure). Active use includes for example messaging or posting content, while passive use includes scrolling. | Systematic review and exploratory meta-analysis |

| | | |
|---|---|---|
| Other | If a measure did not fall under any of the categories above, we classified it as Other. Further, if another category emerged that could be easily quantified in the meta-analysis[1], we discussed with the team whether to include it. | Systematic review and exploratory meta-analysis |

*Note.* [1] No other category emerged that could be classified as separately from the ones defined above.

**Section 3: Inclusion criteria for mental health**

We only included studies that quantitatively measured the mental health outcomes of interest (i.e., anxiety, depression, internalising symptoms). Specifically, we only examined clinical symptom scales of anxiety and depression (i.e., clinical and symptom-based questionnaires on depression/anxiety). In addition, we included scales measuring internalising symptoms if they explicitly reported examining symptoms falling under the realm of depressive and anxiety disorders (including specific types of anxiety such as health anxiety).

**Table 2S.**

Inclusion criteria and examples for the mental health measures.

| Type of mental health outcome | Examples of measures | Use in systematic review/ meta-analysis |
|---|---|---|
| Depression | Beck Depression Inventory (BDI), Center for Epidemiological Studies Depression Scale (CES-D) etc. | Systematic review and confirmatory meta-analysis |
| Anxiety | Revised Child and Anxiety Depression Scale (RCADS), the Generalised Anxiety Disorder scale (GAD-7) etc. | Systematic review and confirmatory meta-analysis |
| Internalising symptoms | Depression Anxiety Stress-Scale (DASS), Hospital Anxiety and Depression Scale (HADS), Mood and Anxiety Symptom Questionnaire (MASQ), Depression Anxiety Stress Scale (DASS), Youth-Pediatric Symptom Checklist (PSC-17) etc. | Systematic review and confirmatory meta-analysis |

**Section 4: Authors contact**

We contacted 18 authors (as denoted in the dataset under the *author_contact* column, dataset available on OSF)[4] for information on the nature of the sample (clinical vs community vs non-clinical) or the conversion of effect sizes. For eight authors, we requested separate coefficients for the relationship between social media use and internalising mental health, based on sample type. Authors from two studies (*ID* = P155, *ID* = P202) replied by providing two separate coefficients for the community/non-clinical and clinical samples.

**Section 5: Conversion of effect sizes to Pearson's r**

Among the selected studies, some reported an effect size other than a Pearson's *r* coefficient (e.g., a Spearman or regression coefficient) to describe the relationship between social media use and internalising symptoms. We included studies that reported a Pearson's *r* or Spearman's *rs* correlation in the article's main text or supplementary materials. For effect sizes initially reported as Spearman's *rs*, we first transformed them to Pearson's *r* and then to Fisher's z. We used the following equation to perform this transformation, $r = 2\sin(rs(\pi/6))$.[5] If a study reported an effect size other than a correlation (e.g., beta regression coefficient or odds ratio), the authors were contacted by email and asked to provide a correlation coefficient. In cases where we received no reply, given that the coefficient provided in the paper was not adjusted (no covariates in the model), we converted coefficients into Pearson's *r*. To convert beta coefficients into *r*, we followed the formula $r = \beta + .05\lambda$.[6] Similarly, we transformed odds ratios into *r* following Lenhard & Lenhard (2016). In total, 17 effect sizes were transformed to Pearson's *r* and were categorised as such in the dataset using the *conversion_r* column ("y", dataset available on OSF).[4]

#### Section 6: Systematic review descriptive results

Below, we report the supplementary results for both the systematic review and meta-analysis.

##### Figure S1

Grid of 10x10 (100%) squares representing the percentage of literature by A) objective (i.e., logged) vs subjective (i.e., self-reported) social media use, B) social media measure, C) global population, D) study design. The presented proportion is calculated based on the total number of effect sizes ($N$ = 775) for A and B, and the total number of studies ($N$ = 127) for C and D.

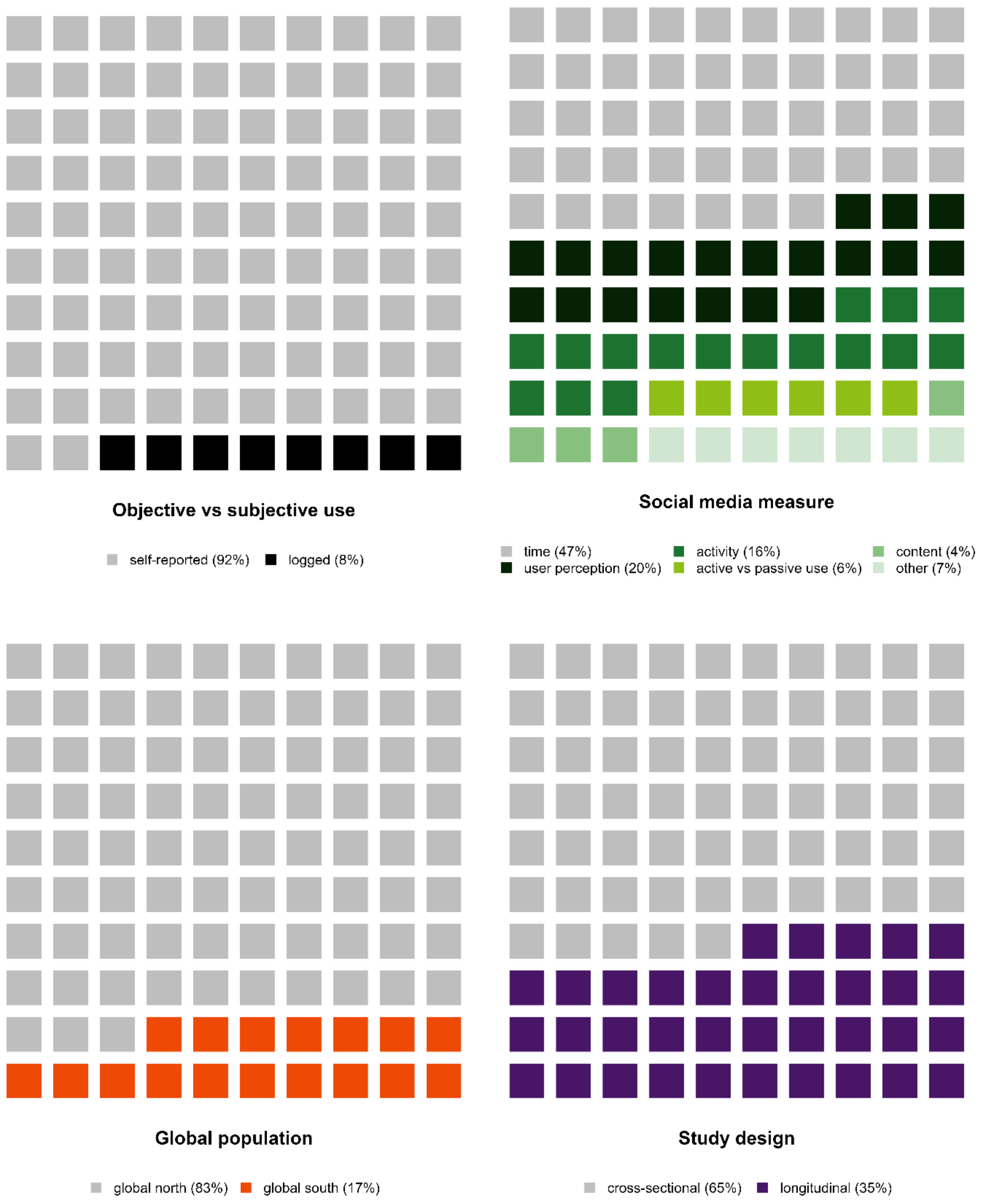

#### Section 7: Funnel plots

##### Figure S2

Observed effect sizes versus their standard errors for comparisons concerning time spent on social media and internalising symptoms in A) all sample types (including both clinical, community and non-clinical), B) all sample types without the identified outlier, C) clinical samples only, D) clinical samples only without the identified outlier, E) community only.

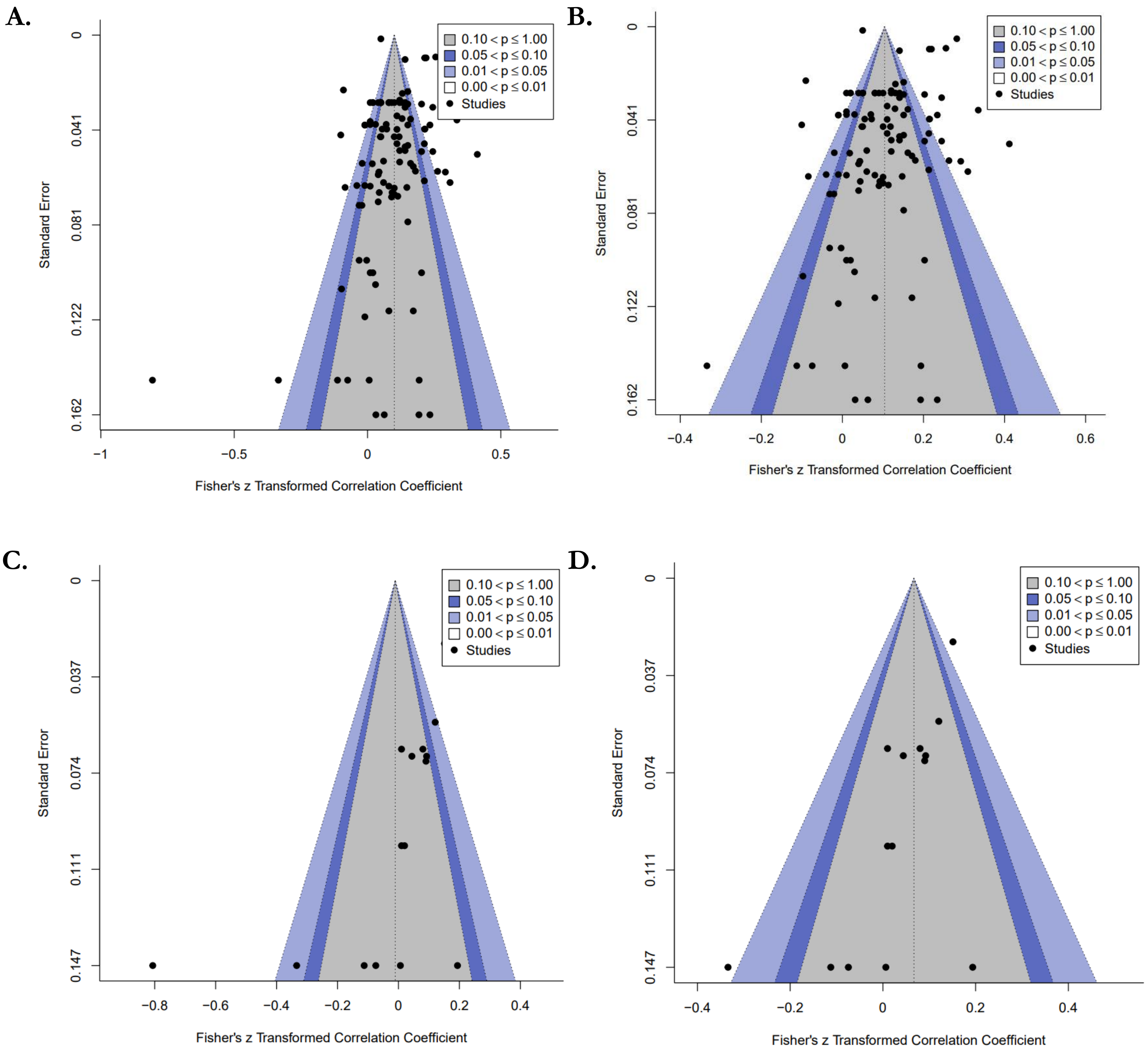

**E.**

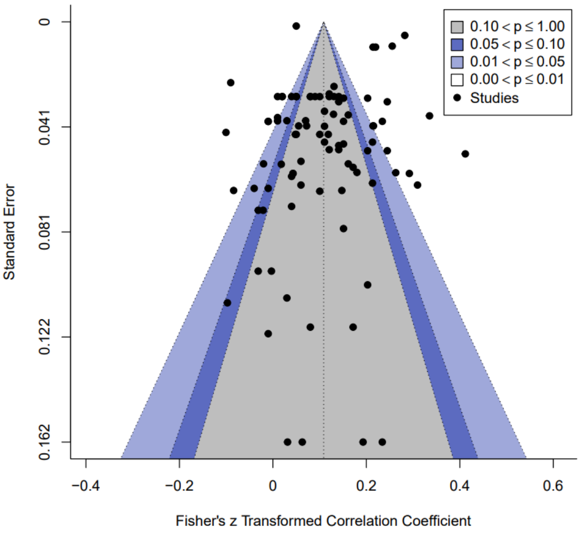


*Note.* The vertical line indicates the estimated summary effect size. The shaded bands represent the significance contours indicated in the legend, and each black dot represents an observed effect size. Visual inspection of all plots does not indicate asymmetry, nor evidence of publication bias as there is no obvious over-representation of effect sizes in the highlighted significance contours. We did not include a funnel plot for the community sample studies without outlier as no outlier was identified based on influence diagnostics.

**Figure S3**

Observed effect sizes versus their standard errors for comparisons concerning social media engagement and internalising symptoms in A) all sample types (including clinical, community and non-clinical), B) all sample types without the identified outlier, C) clinical samples only, D) community only, E) community without the identified outlier.

**A.**

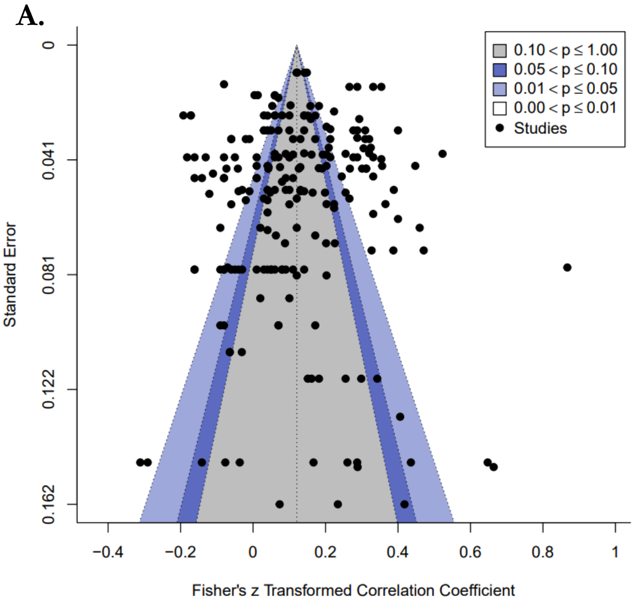


**B.**

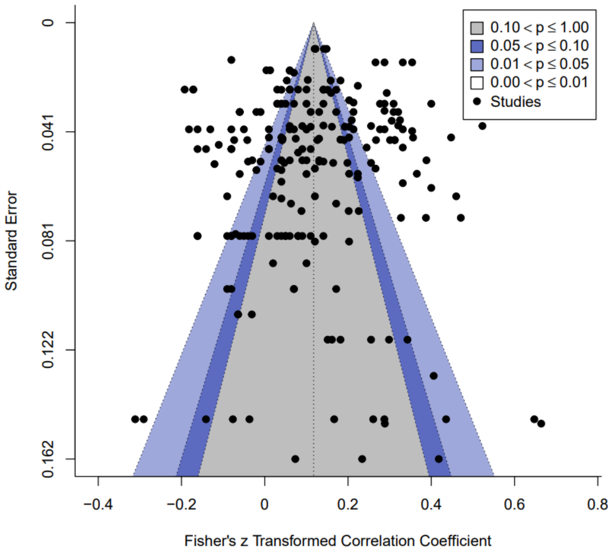

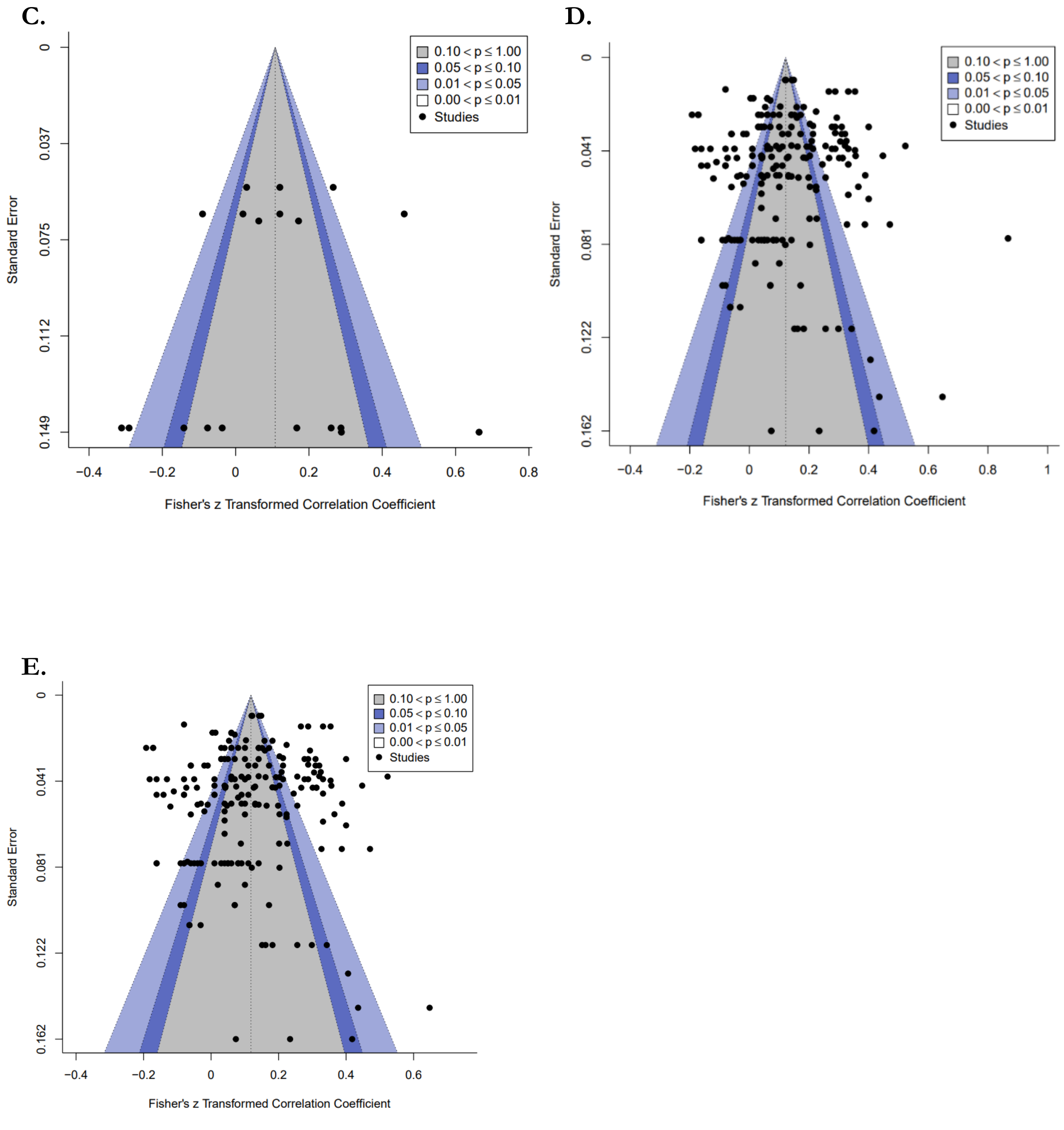


*Note.* The vertical line indicates the estimated summary effect size. The shaded bands represent the significance contours indicated in the legend, and each black dot represents an observed effect size. Visual inspection of all plots does not indicate asymmetry, nor evidence of publication bias as there is no obvious over-representation of effect sizes in the highlighted significance contours. We did not include a funnel plot for the clinical studies without outlier as no outlier was identified based on influence diagnostics.

#### Section 8: Distribution of effect sizes in community samples

**Figure S4**

Caterpillar plot of effect sizes reflecting the correlation in community samples between time spent on social media and internalising symptoms.

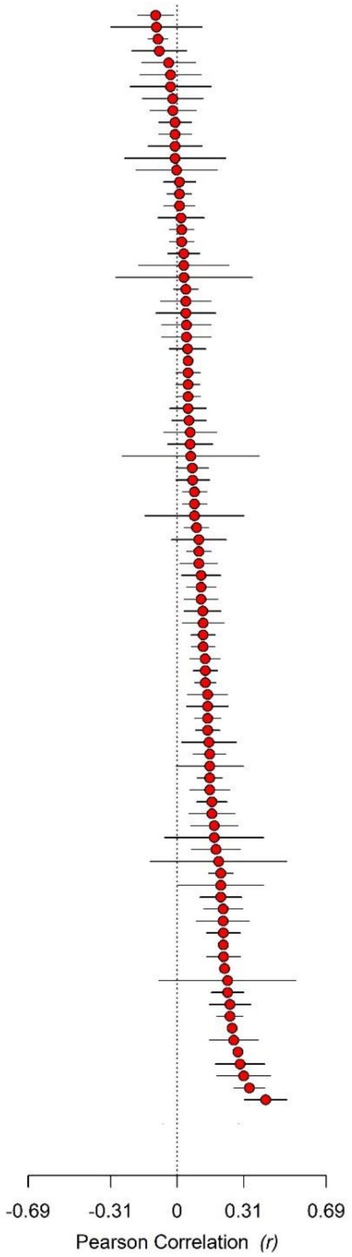


*Note.* Individual Pearson's r coefficients are depicted as filled red circles; the black bars indicate the confidence intervals.

**Figure S5**

Caterpillar plot of effect sizes reflecting the correlation in community samples between social media engagement and internalising symptoms.

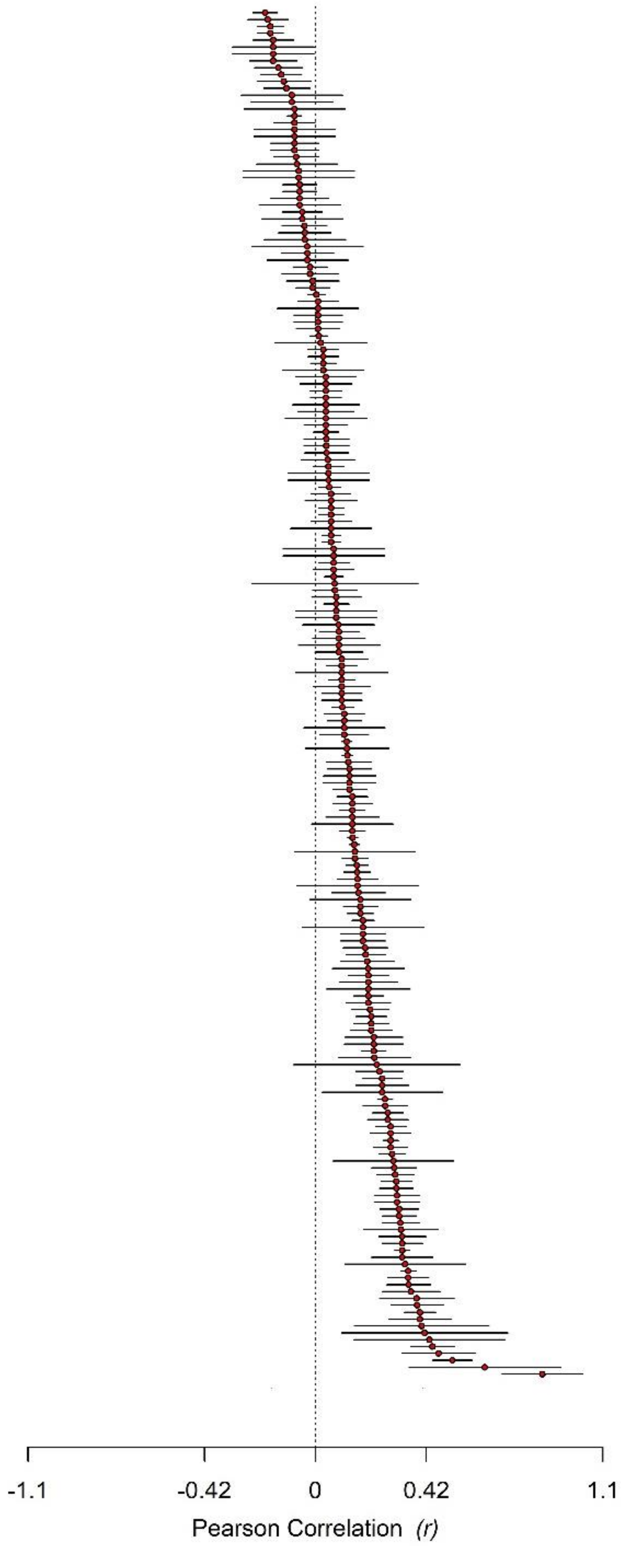


*Note.* Individual Pearson's r coefficients are depicted as filled red circles; the black bars indicate the confidence intervals.

**Section 9: Sensitivity analysis for longitudinal effect sizes**

Our meta-analysis included both cross-sectional and longitudinal studies. Given that longitudinal studies could include multiple waves for the same participants, we only included the effect size from the first wave in the meta-analysis to minimise variance inflation due to dependent effect sizes. Below, we report the sensitivity analyses run to examine changes after including all waves. To summarise, we found no differences in the strength and direction of the meta-analytic effect sizes.

The meta-analysis on the relationship between time spent on social media and internalising symptoms across all sample types included 110 effect sizes (of which 66 were from cross-sectional and 44 from longitudinal studies) from 51 studies. We only included the first wave for longitudinal studies to minimise variance inflation due to dependent effect sizes, adding 3 effect sizes from longitudinal studies with multiple waves. We ran sensitivity analyses to examine whether including all waves would affect the results, leading to 113 effect sizes (66 from cross-sectional and 47 from longitudinal studies) from 52 studies. Our results showed that the inclusion of all waves for time spent on social media did not affect the results from the primary meta-analysis and moderation analysis. The pooled effect size for time spent on social media and internalising symptoms calculated using RVE remained small, positive and significant ($r$ = 0.12, *95% CI* = [0.09, 0.14], $p$ < .001) across all sample types, compared to the primary meta-analysis ($r$ = 0.11, *95% CI* = [0.08 - 0.14], $p$ < .001). Also in this case, sample type was not a significant moderator of the overall relationship between time spent on social media and internalising symptoms ($\beta$ = 0.05, *se* = 0.03, $t$ = 1.7, 95% *CI* = [-0.02, 0.12], $p$ = .153), as was the case in the primary moderation analysis ($\beta$ = 0.05, *se* = 0.03, $t$ = 1.6, 95%*CI* = [-0.02, 0.12], $p$ = .153).

The meta-analysis on the relationship between social media engagement and internalising symptoms in all sample types included 218 effect sizes (of which 162 were from cross-sectional and 56 from longitudinal studies) from 58 studies. We ran sensitivity analyses to examine whether including all waves would affect the results, adding 24 effect sizes from longitudinal studies with multiple waves. This led to 242 effect sizes (66 from cross-sectional and 80 from longitudinal studies) from 58 studies. Our results showed that the inclusion of all waves for social media engagement did not affect the results from the primary meta-analysis and moderation analysis. The pooled effect size for social media engagement and internalising symptoms calculated using RVE remained small, positive and significant ($r$ = 0.15, *95% CI* = [0.12, 0.19], $p$ < .001) across all sample types, compared to the primary meta-analysis ($r$ = 0.15, *95% CI* = [0.12, 0.19], $p$ < .001). Similarly, sample type was not a significant moderator of the overall relationship between time spent on social media and internalising symptoms ($\beta$ = 0.03, *se* = 0.02, $t$ = 1.53, *95%CI* = [-0.03, 0.10], $p$ = .225), as was the case in the primary moderation analysis ($\beta$ = 0.03, *se* = 0.02, $t$ = 1.6, *95% CI* = [-0.03, 0.10], $p$ = .210).

**Section 10: Quality of evidence**

Considering the general shortcomings of study quality assessment in systematic reviews and meta-analyses, such as the lack of focus on empirically verified criteria and the lack of distinction between domains of quality [7] we employed the Quality of Survey Studies in Psychology (Q-SSP) checklist. Details on the rating and applied changes are reported below. Overall, we classified the majority of included papers as acceptable in quality (67/127 studies, 52.6%), with the remainder considered low in quality. Among the 20 items included in the Q-SSP, scores for four items (7 - justification of sample size, 12 - validity of measures, 16 - demographics, 19 - debrief) primarily accounted for lower quality ratings. Altogether, we consider the quality of evidence to be acceptable for our synthesis. Below, we report the quality assessment performed on all studies included in the systematic review, for which we employed the Quality of Survey Studies in Psychology (Q-SSP), with the following changes:

Underlined = Applied changes

1. Was the problem or phenomenon under investigation defined, described, and justified?

2. Was the population under investigation defined, described, and justified?

    - Is adolescence defined, is a community sample clearly defined? Mark as No if they are not.
    - Mark as Yes if some aspect of the sample is defined/justified (e.g. ,"South American adolescents selected due to X" - but no definition of adolescents, would be marked as Yes because the cultural aspect of the population was justified).

3. Were specific research questions and/or hypotheses stated?

4. Were operational definitions of all study variables provided?

5. Were participant inclusion criteria stated?

6. Was the participant recruitment strategy described?

    If study is using existing dataset (e.g., national survey):

    - Mark as No if no link to the sources of data or a protocol published for the dataset is provided.
    - Mark as Yes if a link to the protocol/website of dataset etc. is provided.
    - Mark as Yes if recruitment from schools is stated (no need to state exactly how e.g., email).

7. Was a justification/ rationale for the sample size provided?
    - Mark as NA if authors of the paper did not collect the data.

8. Was the attrition rate provided? (applies to cross-sectional and prospective studies)

    - Mark as Yes for cross-sectional studies that report on excluded cases or participants.

9. Was a method of treating attrition provided? (applies to cross-sectional and prospective studies)
    - Mark as NA if all participants completed study/no data were excluded.
    - Mark No if no mention of attrition or if attrition is reported but no method of treating it (even if it is low).
    - Mark as Yes for cross-sectional studies that have a method of treating missing data (e.g., full information maximum likelihood etc.).

10. Were the data analysis techniques justified (i.e., was the link between hypotheses/ aims / research questions and data analyses explained)?

11. Were the measures provided in the report (or in a supplement) in full?
    - Mark as Yes if clinically validated, open-source or widely used questionnaires are available online.
    - Mark as No if not (i.e., not available online).

12. Was evidence provided for the validity of all the measures (or instrument) used?
    - Mark as No if measures of reliability *only* are provided.
    - For studies that only have one question measuring social media use, consider that measure as valid.
    - Mark as Yes if confirmatory factor analysis (CFA) used to check for construct validity.

13. Was information provided about the person(s) who collected the data (e.g., training, expertise, other demographic characteristics)?
    - Mark as NA if existing dataset used (e.g., national survey).
    - Mark as NA if data collected online.
    - Mark as NA if authors state 'researchers' collected the data.

14. Was information provided about the context (e.g., place) of data collection?
    - Mark as Yes if study is conducted online and this is clearly stated.
    - Mark as No if they have used existing dataset, but not stated the context of data collection (e.g., in school, surveys sent home, online etc).

15. Was information provided about the duration (or start and end date) of data collection?

16. Was the study sample described in terms of key demographic characteristics?
    - Mark as Yes if they include age, gender/sex, race/ethnicity, socioeconomic status (SES) (SES could be a proxy measure such as participant or parent education level etc.).

17. Was discussion of findings confined to the population from which the sample was drawn?
    - Mark as No if no description of sample characteristics provided (e.g., age, gender/sex, country, race/ethnicity, SES) and it is not made clear that the findings are confined to that population.

18. Were participants asked to provide (informed) consent or assent?
19. Were participants debriefed at the end of data collection?

- Mark as NA if debriefing is justifiably waived and an explanation for this provided (e.g., data scraping).

20. Were funding sources or conflicts of interest disclosed?

Additionally:

Various items - mark as Yes if the paper clearly states that the study protocol is described elsewhere and this is accessible (e.g., website, published protocol etc.).

All items - answer in relation to our research questions only (e.g., if authors provide operational definitions of the variables of interest to us, but do not for other variables that are irrelevant to us - mark this as Yes).

**Section 11: Qualitative description of clinical studies**

**Table 3S.**
Key features of the clinical studies included in the systematic review and meta-analysis.

| Authors | Setting | Sample size | Sample description | Design | Social media measure | Mental health measure | Finding | Inclusion |
|---|---|---|---|---|---|---|---|---|
| Akkın Gürbüz et al. (2007)[8] | Global North (Turkey) | N = 108 | Patients were recruited from the Cerrahpaşa Medical Faculty outpatient service, and the control group consisted of high school adolescents who were assumed to have a similar social and demographic background. All patients and controls were evaluated with the K-SADS-PL to diagnose psychopathology. | Cross-sectional | **Time spent, Activity** (via the Social Network Use Questionnaire, measuring online chatting, looking at a friend's activity, sharing a photograph, status update, social games, online games, making friends, video sharing) | **Depressive symptoms:** 10 questions for depressed mood, anhedonia, loss of appetite, insomnia, loss of energy, fatigue, guilt, concentration loss, suicidal ideation and irritability. | Time spent on social media was significantly higher among depressed adolescents than non-depressed adolescents. No differences in social media activities. Additionally, depressed adolescents reported significantly higher disclosure of anhedonia, worthlessness, guilt, loss of concentration, irritability and thoughts of suicide on social media. | Systematic review (excluded from meta-analysis due to lack of coefficients) |
| Fardouly et al. (2020)[9] | Global North (Australia) | N = 49 | Adolescents were recruited via flyers distributed in schools, sports clubs, and medical centres in the city of Sydney, as part of a larger study, the Risks to Adolescent Wellbeing (RAW) Project. Of the 70 (13.3%) participants with a previously diagnosed mental health condition, 49 used social media. The authors provided correlations for the clinical subsample. | Cross-sectional | **Time spent, Activity** (social comparison, appearance investment, selfie posting, likes received). | **Depressive symptoms:** Short Mood and Feelings Questionnaire. | Non-significant correlations between depressive symptoms and social media time spent as well as activities on social media. | Both systematic review and meta-analysis |
| Hamilton et al. (2021)[10] | Global North (USA) | N = 100 | Adolescents enrolled in an intensive outpatient program for depression and suicidality. | Cross-sectional | **Time spent** | **Depressive symptoms:** Short Mood and Feelings Questionnaire. | No overall relationship between social media and depression or average days of suicidal thoughts. Individuals with lower levels of social media use maintained more depression symptoms. | Both systematic review and meta-analysis |
| Kelly et al. (2018)[11] | Global North (UK) | N = 1722 | The Millennium Cohort Study (MCS) is a UK nationally representative prospective cohort study of children born into 19,244 families between September 2000 and January 2002. The authors provided correlations for the clinical subsample. | Cross-sectional | **Time spent** | **Depressive symptoms:** Short Mood and Feelings Questionnaire. | Positive significant correlation between time spent on social media and depressive symptoms. The effect remained after adjusting for family income and structure at age 14, internalising scores at age 11 and age in the regression model. | Both systematic review and meta-analysis |

| | | | | | | | | |
|---|---|---|---|---|---|---|---|---|
| Muzaffar et al. (2018)[12] | Global North (USA) | N = 102 | Adolescents at a suburban safety-net hospital in East Meadow, New York. | Cross-sectional | **Activity** (Facebook behaviours and repetitive behaviours) | **Depressive symptoms:** Mini Mood and Anxiety Symptom Questionnaire; **Social anxiety symptoms**: Leibowitz Social Anxiety Scale for Children and Adolescents, and Anxious Arousal subscale of the Mini Mood and Anxiety Symptom Questionnaire. | Anxiety and depressive symptoms were not associated with Facebook behaviour and repetitive Facebook behaviour. Anxious arousal symptoms were significantly associated with increased Facebook behaviour and increased repetitive Facebook behaviour. | Systematic review (excluded from meta-analysis due to lack of coefficients) |
| Necmi et al. (2018)[13] | Global North (Turkey) | N = 273 | Adolescents that applied to the Uludag Medical Faculty Children and Adolescent Psychiatry Outpatient Clinic, diagnosed with psychopathology based on semi-structured interviews carried out by a child psychiatrist. | Cross-sectional | **Time spent, User perception** (seeking psychological help on social networking sites (SNSs), increased feelings of sociability using SNSs, considering SNSs to be an important part of life, sharing feelings and receiving support on SNSs, feeling more relaxed on SNSs, expressing oneself better on SNSs, share feelings more honestly and openly on SNSs, having supportive friends on SNSs, and emotional disclosure with friends on SNSs) | **Major depressive disorder (MDD) and generalised anxiety (GAD)** - both binary coded - were decided based on a standard semi-structured interview method called Kiddie Schedule for Affective Disorders and Schizophrenia – Present and Lifetime Version (K-SADS-PL). | Non-significant correlation between time spent on social media and MDD or GAD, as well as the perceived value of social media and GAD. Significant positive correlation between MDD and perceived values of social media. | Both systematic review and meta-analysis |
| Nesi et al. (2019)[14] | Global North (USA) | N = 433 | Participants were adolescents who were hospitalized in a psychiatric inpatient facility at an academic medical hospital in the north-eastern United States. | Cross-sectional | **User perception** (A 10-item measure of positive and negative social media experiences was developed based on a review of prior literature) | **Internalising disorder diagnosis:** Children's Interview for Psychiatric Syndromes (ChIPS) to determine psychiatric diagnosis. | Among both boys and girls, youth with internalizing diagnoses were more likely to report having compared themselves negatively to others and having felt excluded on social media compared to adolescents without an internalising diagnosis. | Systematic review (excluded from meta-analysis due to lack of coefficients) |
| Nesi et al. (2021)[15] | Global North (USA) | N = 243 | Participants were adolescents admitted to an adolescent psychiatric inpatient unit. | Cross-sectional | **Time spent, User perception** (perceived overuse, importance of social media, emotional responses to social media experiences) | **Internalising symptoms:** The 17-item version of the Youth-Pediatric Symptom Checklist. | Correlations between time and user perception of social media with internalising symptoms were not significant. | Both systematic review and meta-analysis |
| Nesi et al. (2022)[16] | Global North (USA) | N = 343 | Psychiatrically-hospitalized adolescents. | Cross-sectional | **Time spent, User perception** (impact of social media on positive and negative mood) | **Internalising symptoms:** The 17-item Youth Pediatric Symptom Checklist. | Significant positive correlation of internalising symptoms with both time spent and perceived negative impact of social media. Non-significant correlation between internalising symptoms and perceived positive impact of social media. | Both systematic review and meta-analysis |

| | | | | | | | | |
|---|---|---|---|---|---|---|---|---|
| Noel et al. (2022)[17] | Global North (USA) | N = 212 | Adolescent community sample. The authors provided correlations for the clinical subsample. | Cross-sectional | **Time spent** | **Depressive symptoms**: Center for Epidemiological Studies Short Depression Scale. | Significant positive correlation between depressive symptoms and time spent on social media. | Both systematic review and meta-analysis |
| Twenge et al. (2020)[18] | Global North (UK) | N = NA | The Millennium Cohort Study (MCS) is a UK nationally representative prospective cohort study of children born into 19,244 families between September 2000 and January 2002. Subsample based on depressive symptoms above clinical cut-off. | Cross-sectional | **Time spent** | **Depressive symptoms**: Short Version of the Mood and Feelings Questionnaire. | Association between time spent on social media and clinically relevant depressive symptoms. | Systematic review (excluded from meta-analysis due to lack of coefficient and sample size of the clinical subgroup) |
